# Physician Responses to Apple Watch-Detected Irregular Rhythm Alerts: A Case-Based Survey

**DOI:** 10.1101/2022.08.02.22278237

**Authors:** Patrick C. Demkowicz, Sanket S. Dhruva, Erica S. Spatz, Alexis L. Beatty, Joseph S. Ross, Rohan Khera

## Abstract

**Background:** While the US Food and Drug Administration (FDA) has cleared smart watch software for detecting atrial fibrillation (AF), there is lack of guidance on management by physicians.

**Methods:** We conducted a case-based survey of physicians in primary care, emergency medicine, and cardiology at Yale and University of California San Francisco from September to December 2021. Cases described asymptomatic patients receiving Apple Watch AF alerts; cases varied in sex, race, medical (diabetes and hypertension) history, and notification frequency. Physicians were asked to select from among prespecified diagnostic testing, referral, and treatment options.

**Results:** We emailed 636 physicians, of whom 95 (14.9%) filled out the survey, including 39 primary care, 25 emergency medicine, and 31 cardiology physicians. Among a total of 192 cases (16 unique scenarios), physicians selected at least one diagnostic test in 191 (99.5%) cases and medications in 48 (25.0%). Rates of patient referral (14%, 30%, and 16%, respectively; P=.048), rhythm monitoring (84%, 46%, and 94%, respectively; P<.001), measurement of BNP (8%, 20%, and 2%; P=.003), and use of antiarrhythmics (16%, 4%, and 23%; P=.023) varied among primary care, emergency medicine, and cardiology, respectively. Management was largely consistent across case characteristics (race, sex, medical history, and alert frequency).

**Conclusion:** In hypothetical cases of patients presenting without clinical symptoms, physicians opted for further diagnostic testing and often to medical intervention based on Apple Watch irregular rhythm notifications. There was also considerable variation across physician specialties, suggesting a need for uniform clinical practice guidelines.

## BACKGROUND

Several medical and public health bodies do not recommend widespread screening for atrial fibrillation (AF). The US Preventative Services Task Force determined in 2022 that current evidence is insufficient to determine balance of benefits and harms of ECG screening among asymptomatic adults.^1^ Screening for AF has not been demonstrated to reduce stroke risk.^2-4^ Conversely, healthy patients who falsely screen positive may receive unnecessary and potentially harmful testing and treatment.^5,6^ Despite such evidence, there is widespread interest in screening for AF with the goal of reducing stroke mortality and morbidity. The FDA has cleared smart watch applications that can identify irregular rhythms, such as AF, regardless of whether the patient experiences clinical symptoms, including recent versions of the Apple Watch and Fitbit.^7,8^ Some devices also have single-lead ECG functionalities that clinicians may manually review. However, there is no evidence that medical evaluation for irregular rhythms detected by smart watches improves patient morbidity or mortality.

Despite a lack of established benefits of wearable screening for AF, smart watches have grown in popularity with 1% of primary care patients having documentation of a device in a recent study of an academic health care system.^9^ Clinical encounters where patients discuss information generated from smart watches are challenging for physicians given the lack of clinical evidence to inform decision-making, clinical practice guidelines regarding asymptomatic AF screening, and consensus strategies for clinical evaluation for these patients. Furthermore, different device companies’ algorithms vary in their diagnostic yield.^10^ Recent surveys have found that cardiac electrophysiologists would consider pursuing further electrophysiological evaluation as well as initiate anticoagulation in asymptomatic patients on the basis of smart watch irregular rhythm and single-lead ECG findings despite lack of any evidence to suggest benefit of such an approach.^11,12^ These findings suggest that there is substantial variation in the strategies adopted by different clinicians.

On the other hand, ssssssswidespread availability of smart watches may instead increase rates of AF diagnosis among marginalized patients, mitigating current disparities in care. For example, Black patients with AF are less likely than White patients to receive a formal diagnosis and receive anticoagulation.^13^ Female patients with AF are less often prescribed anticoagulation therapy despite having higher stroke risk.^14^ Furthermore, a study using hypothetical case histories found gender bias in attitude toward and secondary prevention of patients with coronary artery disease.^15^ A study using actors portraying patients with chest pain found that women and Black adults were less likely to be referred for cardiac catheterization than men and White adults.^16^

In this study, we used hypothetical patient cases to conduct a multicenter evaluation of physicians’ responses to asymptomatic patients reporting Apple Watch irregular rhythm notifications suggestive of AF. We sought to evaluate variation across physicians and assess whether specialty and personal experience with smart watches influenced their approach to management. We also assessed whether patient characteristics influenced the approach of physicians to patient-reported episodes of Apple Watch-detected, asymptomatic AF.

## METHODS

### Study Population

We conducted a cross-sectional, case-based survey of attending and resident physicians practicing in primary care, emergency medicine, and cardiology departments at two geographically separated health systems, (1) Yale New Haven Hospital in New Haven, Connecticut and, (2) University of California San Francisco between September and December 2021. Respondents’ department affiliations and email addresses were gathered from their institutional directory profiles, if available. Our study was exempted from review by the Yale and UCSF institutional review boards.

### Survey Design

The survey was developed by 2 cardiologists, an internist, and a medical student, and reviewed by 2 other cardiologists. The survey is included in Supplement 1 and the Consensus-Based Checklist for Reporting of Survey Studies (CROSS) is included in Supplement 2.^17^ The survey consisted of two clinical vignettes. Each vignette involved a 60-year-old person who reported recently receiving one or more irregular rhythm notifications on their Apple Watch in the absence of associated symptoms (i.e., no fatigue or racing sensation in his chest). Four variables varied across the 16 scenarios: stroke risk-factors (none or history of diabetes and hypertension), reported frequency of recent alerts (1 or >1), sex (male or female) and race (Black or White). The atrial fibrillation stroke risk factors represent items on the CHAD_2_DS_2_-VASc score.^18^

Respondents were randomized by the survey platform to see one low stroke-risk case and one moderate-high stroke-risk case of two patients with the same race and sex. Due to an error in the survey platform settings, 5 respondents completed a third case that varied from their second case with respect to alert frequency. A respondent notified the team of the error and it was promptly corrected. Respondents were then asked to rate on a five-point Likert scale the likelihood that they would consider specific diagnostic evaluations and therapeutic interventions. Next, respondents were asked on a five-point Likert scale how important (extremely, very moderately, slightly, not at all) various factors were in determining their answer choices in the preceding cases: likelihood of AF in patient’s group, risk of stroke in patient’s group, strength of evidence, concern about missing a diagnosis, and concern about unnecessary testing. Finally, respondents were asked about their own backgrounds and practices. We asked respondents about their gender, race, specialty, training history, personal use of smart watches capable of rhythm detection, and experience recommending smart watches to their patients.

### Survey Delivery

We used Qualtrics to host our survey and send email invitations. Qualtrics generated a unique invitation link for each respondent. This allowed us to track survey completion and prevent multiple participation. Completed responses were deidentified by Qualtrics. Respondents at Yale were emailed three weekly reminders and respondents at UCSF 2 reminders (1 and 3 weeks) if they did not respond. Respondents were shown an information sheet about the survey before consenting to participate. They were then offered the opportunity to enter a drawing for a $100 gift card. To begin the survey, respondents were required to consent to participate and then select their specialty before they could advance to the case scenarios. Respondents who did not complete this step or who left all case questions unanswered were excluded.

### Outcomes

The primary outcome was the proportion of responses indicating the use of specific interventions. These included: referring to primary care, cardiology, or electrophysiology; ordering a cardiovascular stress test, 12-lead ECG, serum brain natriuretic peptide (BNP), transthoracic echocardiogram, ambulatory rhythm monitoring (specific options included event monitor, implantable loop recorder, patch monitor, or a commercially available heart rhythm monitor such as AliveCor, which were combined into one category); and treating with aspirin, anticoagulation, a beta blocker or calcium channel blocker, or a class IC or III antiarrhythmic. We categorized the responses as “yes” and “no”, combining responses of “Extremely Likely” or “Somewhat Likely” into a yes category and responses of “Neither Likely nor Unlikely,” “Somewhat Unlikely,” and “Extremely Unlikely” into a no category.

### Statistical Analyses

We generated descriptive statistics (specialty, sex, age, and years in practice) about our study population using percentages to report categorical variables and averages (standard deviation) to report continuous variables, stratified by respondent specialty. We also reported rates of different categories of experiences with smart watches (personal use, having previously recommended patients use wearable devices, and having experienced a patient reporting results from a smart watch) stratified by respondent specialty. We used the chi-squared test to compare rates across specialties.

We reported the rates at which specific tests and interventions were selected. Because respondents evaluated multiple cases, we calculated mean rates out of all, non-unique cases. We also used a histogram distribution to report the number of diagnostic tests (stress test, 12-lead ECG, serum BNP, TTE, ambulatory rhythm monitoring), or medical interventions (aspirin, anticoagulation, antiarrhythmic) that were selected across all, non-unique cases.

We analyzed the relationship between respondent characteristics and the likelihood with which specific tests or interventions were selected. We repeated this analysis to compare across years of practice (0-9, 10-19, 20+) and between respondents who did or did not report different types of experiences with smart watches. We used the chi-squared test to compare rates across levels of respondent characteristics (e.g., specialty).

We repeated the above analysis to evaluate the relationship between patient characteristics and respondent’s likelihood of selecting each test or intervention. We compared rates of each intervention by the race (Black vs. white), sex (male vs. female), and history of stroke risk factors (none vs. hypertension and diabetes) of the patient described, as well as by the number of notifications reported by the patient in the scenario (one vs. many). Analyses were conducted using R (version 4) and figures were generated using Prism (version 9). A 2-sided p value <0.05 was considered statistically significant.

## RESULTS

### Study Population

We emailed 636 physicians (excluding an additional 14 emails that bounced-back or failed), of whom 95 (14.9%) completed the survey, including 39 primary care, 25 emergency medicine, and 31 cardiology physicians (Table 1); 75 were based at Yale and 20 at UCSF. Overall, 44.2% of respondents identified as female, 55.8% were aged 44 years or younger, and 40% of respondents report being practice less than a decade, with 22.1% practicing 10-19 years, and 31.6% practicing more than 20 years (6.3% missing).

**Table 1.**
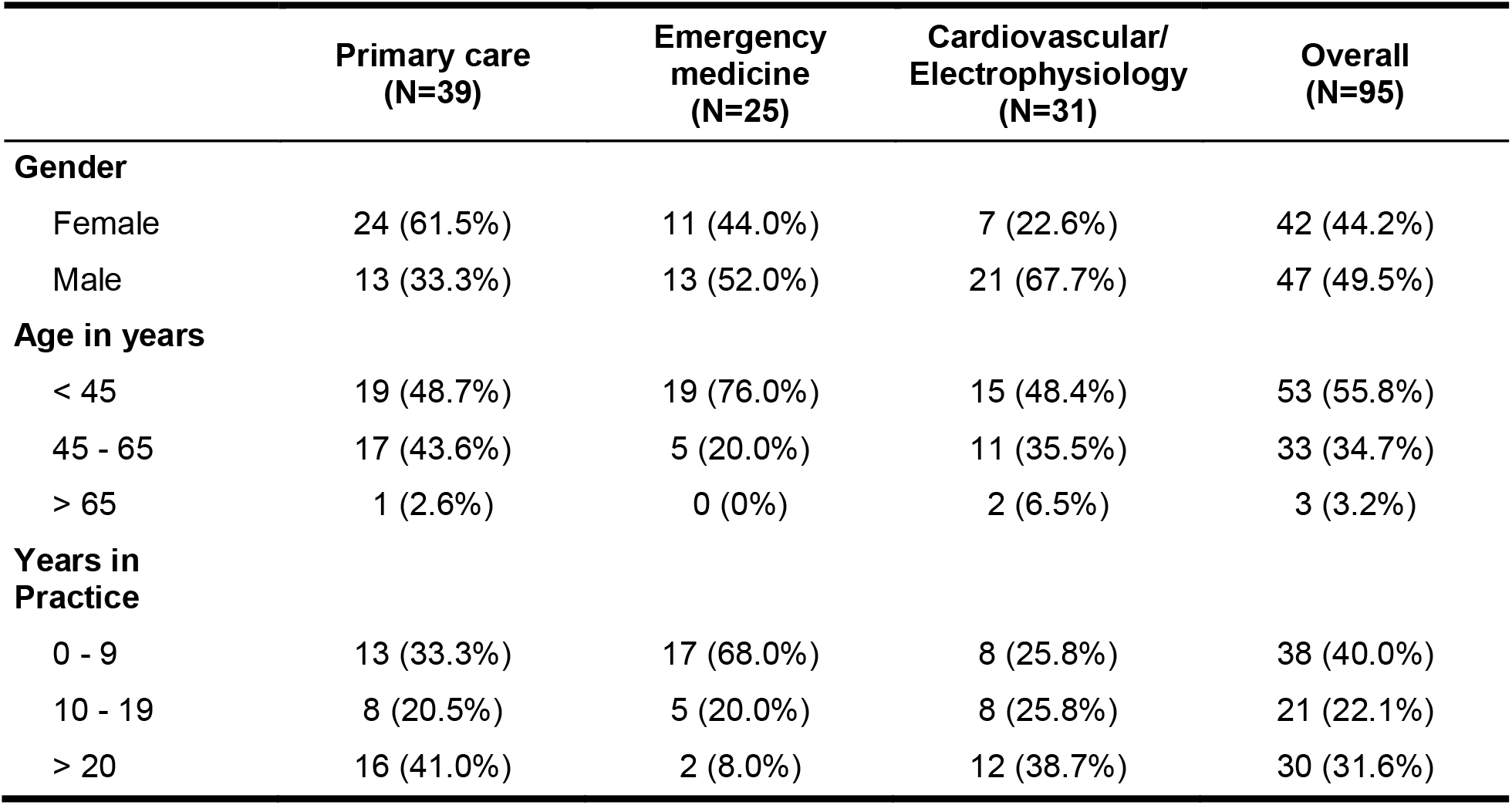
Respondent characteristics. Respondent demographics and years in practice are presented overall and by specialty. Data were missing for 6 respondents (6.3% overall).

|  | Primary care<br>(N=39) | Emergency<br>medicine<br>(N=25) | Cardiovascular/<br>Electrophysiology<br>(N=31) | Overall<br>(N=95) |
| --- | --- | --- | --- | --- |
| **Gender** |  |  |  |  |
| Female | 24 (61.5%) | 11 (44.0%) | 7 (22.6%) | 42 (44.2%) |
| Male | 13 (33.3%) | 13 (52.0%) | 21 (67.7%) | 47 (49.5%) |
| **Age in years** |  |  |  |  |
| < 45 | 19 (48.7%) | 19 (76.0%) | 15 (48.4%) | 53 (55.8%) |
| 45 - 65 | 17 (43.6%) | 5 (20.0%) | 11 (35.5%) | 33 (34.7%) |
| > 65 | 1 (2.6%) | 0 (0%) | 2 (6.5%) | 3 (3.2%) |
| **Years in**<br>**Practice** |  |  |  |  |
| 0 - 9 | 13 (33.3%) | 17 (68.0%) | 8 (25.8%) | 38 (40.0%) |
| 10 - 19 | 8 (20.5%) | 5 (20.0%) | 8 (25.8%) | 21 (22.1%) |
| > 20 | 16 (41.0%) | 2 (8.0%) | 12 (38.7%) | 30 (31.6%) |

Overall, 27 (28.4%) respondents reported personally using a smart watch, which was consistent across specialties (P=.68) (Table 2). However, respondents of different specialties reported considerably different experiences with smart watches in their clinical practices, including the rates at which they had recommended smartwatches to their patients (12.8% primary care, 12.0% emergency, and 41.9% cardiology; P=.003) and have had a patient report a smart watch alert (30.8%, 80.0%, and 80.6%, respectively; P<.001).

**Table 2.** Experience with smartwatches. Respondents’ experiences with smartwatches (personally wearing one, having had recommended one to a patient, and having encountered a patient reporting smartwatch alerts) are presented by specialty. Data were missing for 6 respondents (6.3% overall).

|  | Primary care<br>(N=39) | Emergency<br>medicine<br>(N=25) | Cardiovascular/<br>EP<br>(N=31) | P-value |
| --- | --- | --- | --- | --- |
| **Wears a Smartwatch** |  |  |  | 0.681 |
| Yes | 13 (33.3%) | 6 (24.0%) | 8 (25.8%) |  |
| No | 24 (61.5%) | 18 (72.0%) | 20 (64.5%) |  |
| **Has Recommended Smartwatches to Patients** |  |  |  | 0.003 |
| Yes | 5 (12.8%) | 3 (12.0%) | 13 (41.9%) |  |
| No | 32 (82.1%) | 21 (84.0%) | 15 (48.4%) |  |
| **Has Experience with Patients Reporting Smartwatch Alerts** |  |  |  | <0.001 |
| Yes | 12 (30.8%) | 20 (80.0%) | 25 (80.6%) |  |
| No | 25 (64.1%) | 4 (16.0%) | 3 (9.7%) |  |

### Overall Approach to Case Scenarios

Respondents completed a total of 192 cases drawn from 16 unique scenarios; 3 respondents completed 1 case, 87 respondents completed 2 cases, and 5 respondents completed 3 cases. In 191 (99.5%) cases physicians selected at least one diagnostic test to work-up the asymptomatic Apple Watch irregular rhythm notification: electrocardiography (185, 96.4%), ambulatory rhythm monitoring (148, 77.1%), transthoracic echocardiography (TTE) (63,

32.8%), stress testing (19, 9.9%), and brain natriuretic peptide (BNP) evaluation (17, 8.9%) (Figure 1). Respondents selected an average of 2.3 diagnostic tests (standard deviation 0.97) (Figure 2A). In addition, referral and treatment options were commonly selected in response to the notification: in 36 (18.8%) cases physicians selected referral to a different specialty and in 48 (25.0%) new medication treatment, such as aspirin (35, 18.2%), antiarrhythmics (29, 15.1%), and/or anticoagulation (18, 9.4%). Respondents selected an average of 0.4 types of medication (standard deviation 0.8) (Figure 2B). When asked about the factors that influenced their answers to the cases, 50.5%, 42.1%, 56.8%, 43.2%, and 48.4% of respondents rated as very important or extremely important the likelihood of AF, the risk of stroke, the strength of evidence, concern about missing a diagnosis, and concern about unnecessary testing, respectively (Supplement 3, Table 1)

**Figure 1.**
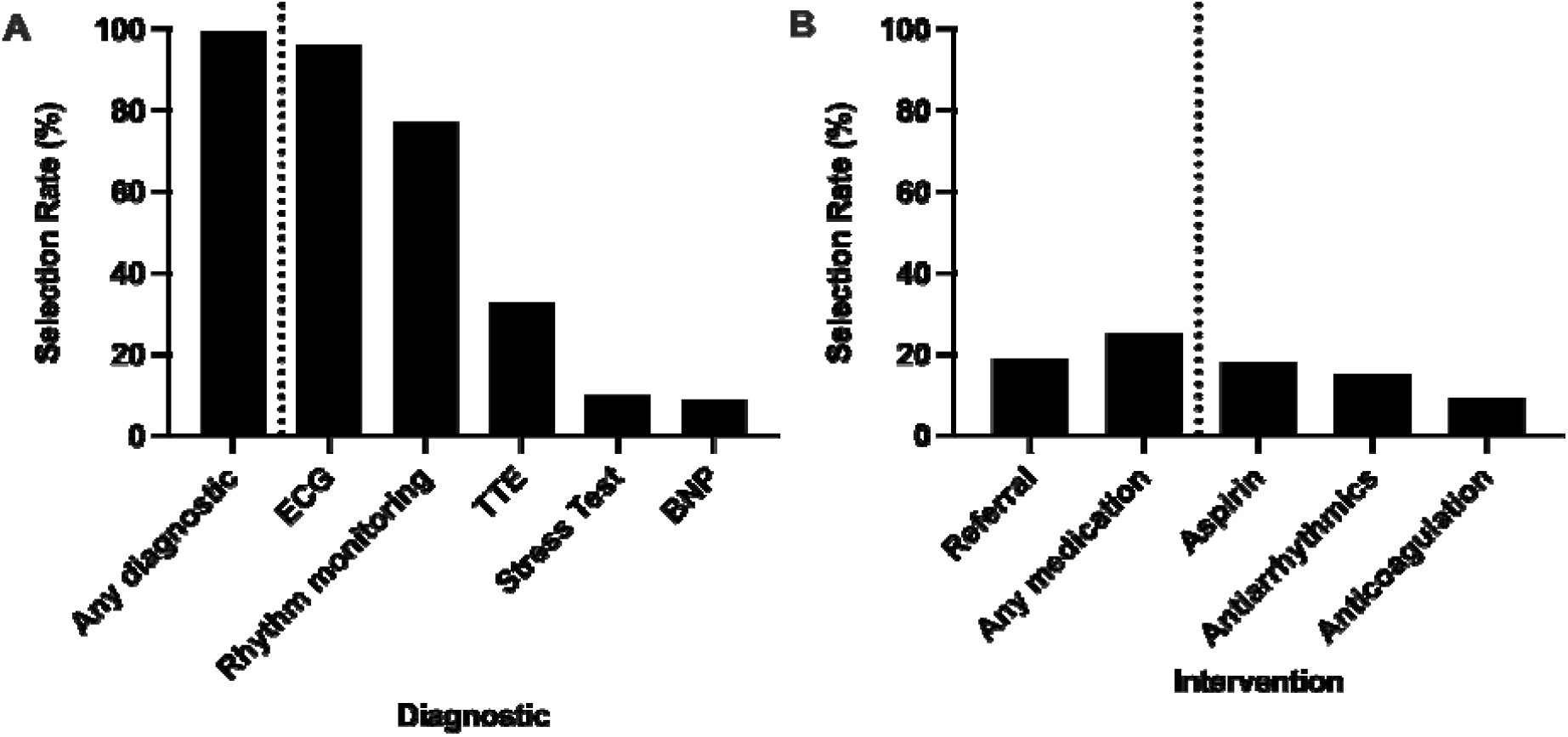
Rates of testing and intervention overall. Among 95 participants, 192 hypothetical cases were completed. Here we report the average rate at which respondents indicated that they were “extremely likely” or “somewhat likely” to order specific diagnostic tests, referral/consultation, and therapeutic interventions. “Rhythm monitoring” indicates that any of the following were selected: event monitor, implantable loop recorder, patch monitor, or a commercially available heart rhythm monitor such as AliveCor. “Antiarrhythmics” included selection of a beta blocker or calcium channel blocker, or a class IC or III antiarrhythmic. ECG = electrocardiogram, TTE = transthoracic echocardiogram, and BNP = brain natriuretic peptide.

**Figure 2.**
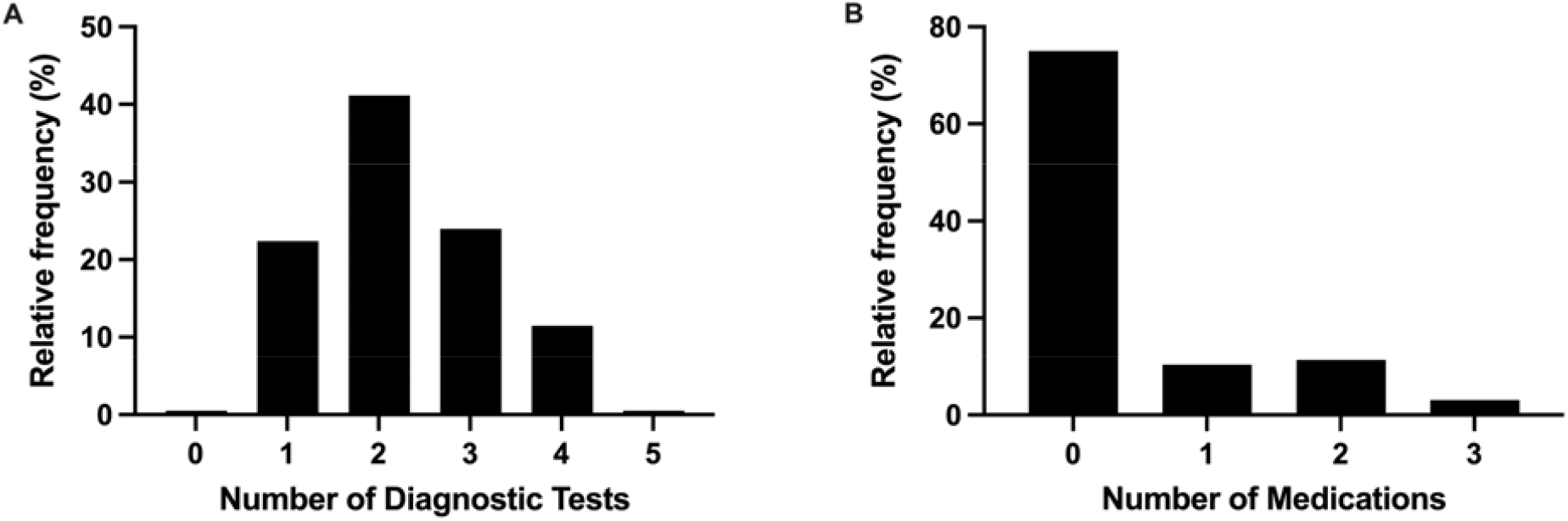
Frequency of (A) diagnostic testing and (B) medication treatment across cases. The number of diagnostic tests (among ECG, rhythm monitoring, TTE, stress test, and BNP) and types of medications (among aspirin, antiarrhythmics, and anticoagulation) were tallied in each case (N = 192). The distributions of frequencies are reported here.

### Clinician Factors and Approach to Case Scenarios

With regard to diagnostic testing, there were no statistically significant differences among specialties in selection of ECG, TTE, or stress testing (Figure 3A). With regard to interventions, there were no statistically significant differences among specialties in selection of aspirin or anticoagulation (Figure 3B). There were significant differences among specialties in rates of referral (P=.048), BNP measurement (P=.003), ambulatory rhythm monitoring (P<.001) and prescription of antiarrhythmic therapy (P=.023). Years spent in practice was associated with use of transthoracic echocardiogram (P=.023) and ambulatory rhythm monitoring (P<.001) (Supplement 3, Figure 1).

**Figure 3.**
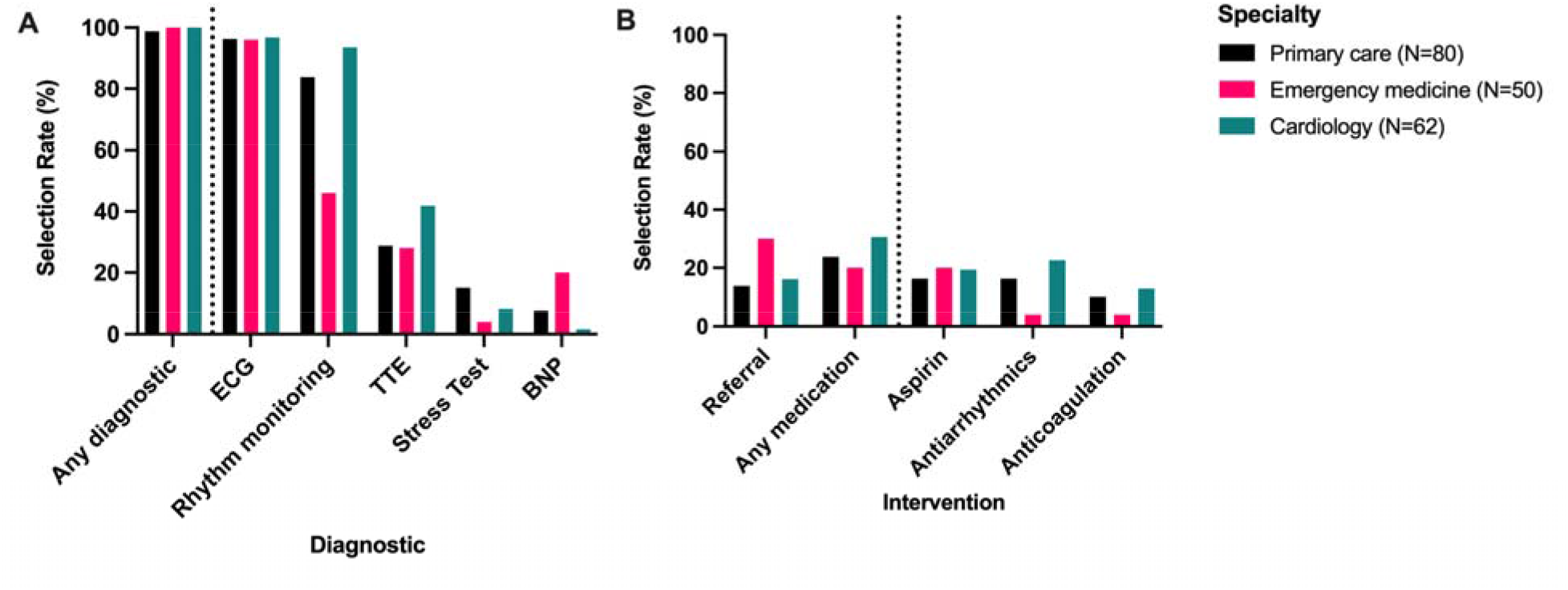
Rates of testing and intervention by respondent specialty.

Respondents who have previously recommended their patients to use smart watches were more likely to order TTEs (54.8% vs. 24.8%, P<.001) and ambulatory rhythm monitoring (95.2 vs. 70.2%, P=.002) (Supplement 3, Table 2). There was no relationship between other respondent factors, including wearing a smart watch, having treated patients who reported smart watch findings, and having recommended smart watches to patients, and selection of specific interventions.

### Simulated Patient Factors and Overall Approach to Case Scenarios

The distribution of case characteristics is summarized in Supplement 3, Table 3. Black race was associated with a higher receipt of antiarrhythmics (21.3% vs. 9.2%; P=.033) (Figure 4). Selection of other diagnostics and interventions were otherwise similar between cases describing Black and White patients. Female sex was associated with lower use of serum BNP (4.0% vs. 14.1%; P=.027). Selection of other diagnostics and interventions were otherwise similar between cases describing female and male patients. Patient stroke risk (presence vs. absence of diabetes and hypertension) and reported frequency of alerts (single vs. repeated) were not associated with differential intervention rates (Supplement 3, Table 4).

**Figure 4.**
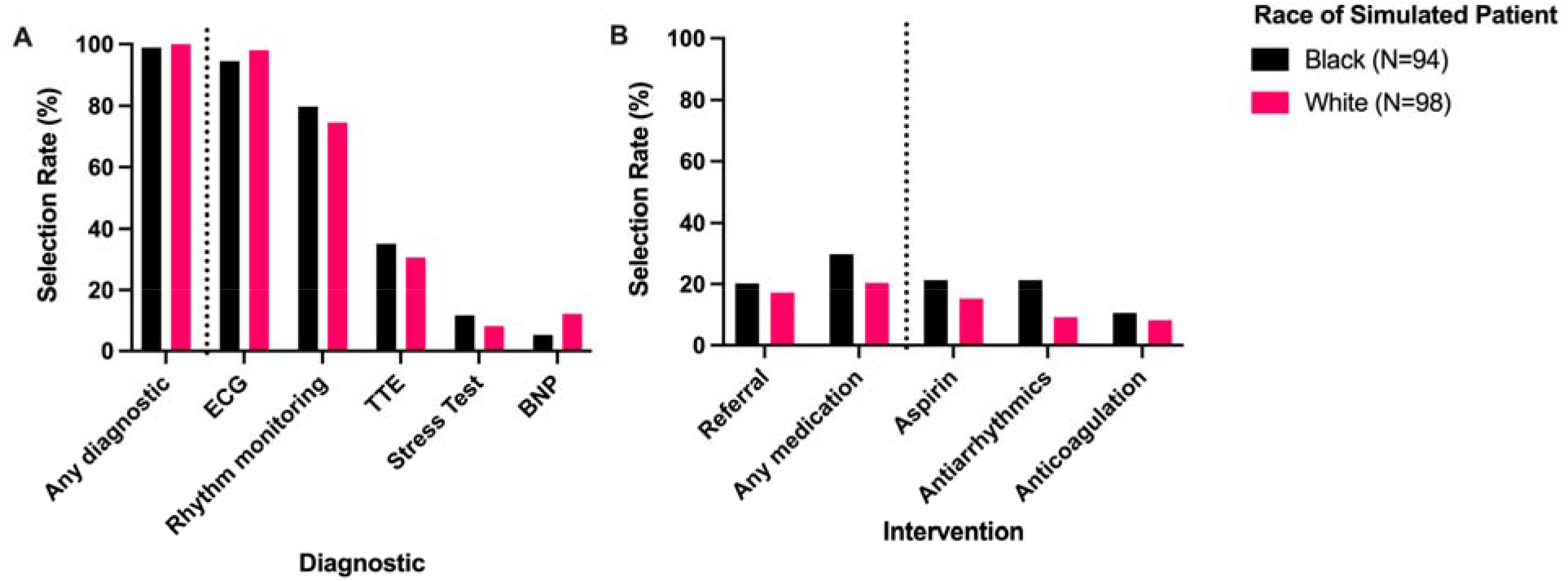
Rates of testing and intervention by race of simulated patient.

## DISCUSSION

In this survey of physicians at two institutions, we used hypothetical clinical vignettes to examine decision-making patterns in response to irregular rhythm notifications on an Apple Watch in patients without symptoms of AF. Notifications nearly always triggered further diagnostic workup, varying from low-cost and low-intensity tests such as ECG to higher-cost and higher-intensity procedures such as stress testing. Furthermore, in 1 out of 4 simulated encounters, respondents considered initiating medications, including antiplatelet, anticoagulant, and antiarrhythmic therapies on initial presentation. We found that management did not vary substantially across case characteristics such as patient race, sex, stroke risk, and alert frequency.

Our findings are consistent with prior studies that have examined clinical decision-making based on rhythm data from smart watches. A 4-month single-center retrospective review identified 264 patients evaluated for abnormal pulse detected using Apple Watch, 33% of whom were asymptomatic.^19^ The study found that 61% of asymptomatic patients underwent diagnostic testing. The study also found variation in diagnostic testing across clinical departments, with patients seen in the emergency department more likely to undergo 12-lead ECG or bloodwork compared with patients seen in primary care or by a cardiologist. One possible explanation for a higher rate of diagnostic testing in our survey is that respondents were shown only brief vignettes and not afforded the opportunity to perform a physical examination. In the retrospective study, clinicians had preexisting relationships with many patients: nearly half of all patients had a preexisting cardiovascular diagnosis and the most common department of presentation was cardiology. Clinicians may therefore be able to work up smart watch irregular rhythm notifications more conservatively in the real world than in our survey because they have more data to inform decision-making.

Prior studies examining prescribing patterns in response to smart watch notifications have focused on the interpretation of single-lead ECG tracings available on models such as the Apple Watch 4 and later. A survey of 1601 clinicians, including advanced practice providers, found that results from a 30-second single-lead ECG were sufficient for 42.7% of clinicians to recommend oral anticoagulation for patients at high risk for stroke.^11^ A survey of 417 electrophysiologists worldwide found that, when presented with a single-lead ECG tracing suggesting AF, 21% would consider initiating anticoagulation in an asymptomatic patient.^12^ In contrast, we found that respondents would consider anticoagulation approximately 9% of the time in response to irregular rhythm notifications. It is expected that clinicians would be less likely to consider anticoagulation on the basis of an irregular rhythm notification than a single-lead ECG tracing that they can manually review. Manual clinician review of single-lead ECG tracings can improve the diagnostic utility of the Apple Watch automated AF-detection algorithm.^10^ However, it is not reasonable to initiate anticoagulation solely after a notification because prior studies of Apple Heart Study participants found that follow-up ambulatory ECG confirmed AF in 34% of cases of irregular rhythm notification, with non-AF irregular rhythms detected in 40% of cases without AF.^20,21^

It is notable that stroke risk in our study was not associated with respondent practice patterns. Researchers have advocated for screening for AF in the high-stroke risk population with the assumption that such patients would have a greater benefit.^22^ Our findings show that physicians approached patients with the least potential benefit from AF screening and treatment similarly to how they approached patients who would have the greatest potential benefit. This suggests that in actual practice, physicians might apply such screening guidelines with a much larger scope that recommended. This finding supports the concern that smart watch utilization may lead to greater health care expenditure on potentially unnecessary testing and treatment.^23^ In patients who have true paroxysmal AF, this may also lead to disparities in access to cardiovascular care between patients who own smart watches and patients who do not.

It is also notable that, in contrast with prior studies using simulated patients, there was little variation in management with respect to the patient’s race or sex.^15,16^ This study had fewer respondents and therefore may not have been sufficiently powered. Furthermore, our respondent population skewed toward younger clinicians, who may have had greater exposure to anti-bias curricula in their medical training than clinicians later than their careers.^24^ This finding also only looks at one mechanism – clinician bas – of systemic racism and sexism, which may affect the outcomes of real-world patients in a myriad of ways.^25^ Further research is needed to understand disparities in the treatment of smart watch-detected irregular rhythms in real world clinical practice.

There are several limitations to our study. First, 14.9% of physicians contacted ultimately responded to our survey; respondents may have stronger attitudes toward smart watches than non-respondents. We found in our survey that respondents with positive attitudes toward smart watches were more likely to select certain interventions. Second, we assessed practice patterns using written clinical case vignettes. It is assumed that respondents would act similarly when managing actual patients.^16^ Furthermore, assessments made on the bases of written case vignettes have been shown to correlate with those made on the basis of in-person examinations. Third, our case scenarios were limited in detail and other potentially informative clinical cues available in the real-world were not provided. Lastly, we surveyed physicians at two large academic centers, which may limit the generalizability of our findings.

## CONCLUSIONS

Our survey demonstrates that many physicians likely have a high degree of confidence in smart watch irregular rhythm notifications as demonstrated by respondents’ likelihood of pursuing additional diagnostic testing and interventions. This finding raises several concerns: smart watch utilization may lead to greater health care expenditure on potentially unnecessary testing and treatment; it may also lead to disparities in access to cardiovascular care between patients who own smart watches and patients who do not. Therefore, despite the lack of recommendation by public health and professional organizations for AF screening in asymptomatic patients, our study highlights the need for further evidence to inform the development of standardized guidelines.

## Data Availability

All data produced in the present study are available upon reasonable request to the authors.

## Author Contributions

All authors were responsible for study concept and design; PD, RK, and SD for acquisition of subjects and data; PD, RK, and SD, and JR for analysis and interpretation of data; and all authors for preparation of manuscript.

