## Supplement 1 for "Physician Responses to Apple Watch-Detected Irregular Rhythm Alerts: A Case-Based Survey"

Wearable device survey

Start of Block: Disclaimer

[INFORMATION SHEET FOR PARTICIPATION IN A RESEARCH STUDY]

Do you agree to participate?

- Yes
- No

End of Block: Disclaimer

Start of Block: Specialty Screen

Thank you for volunteering to participate in this survey. To begin, please select the answer choice that best describes your most advanced level of training (residents and fellows should select the training they are currently in the process of completing):

- Primary care (including family medicine)
- Emergency medicine
- Cardiovascular (not electrophysiology)
- Electrophysiology

End of Block: Specialty Screen

Start of Block: Instructions for Part I

**Part I of II**
 In this section, you will be shown two clinical scenarios, each describing a patient visiting your clinic.  After reviewing the details of the patient presentation, you will be asked to indicate how likely you would order a series of evaluations, ordered alphabetically.  Please check all that apply.

End of Block: Instructions for Part I

B.M.0.S

A 60-year-old black male with no pertinent past medical history presents after receiving a notification on his Apple Watch this past week that his heart showed “signs of an irregular rhythm suggestive of atrial fibrillation.” He explains that he ignored the notification because he thought something was wrong with the watch, but figured he should get checked out. The patient denies symptoms of palpitations, dyspnea or fatigue. Vital signs reveal a temperature of 37.0, blood pressure of 120/80, heart rate of 65, respiratory rate of 12, and BMI of 29. Physical exam does not reveal irregular pulse, irregular jugular venous pulsations, or abnormal heart sounds. He does not smoke tobacco and drinks alcohol only occasionally, at social events.

B.M.0.R

A 60-year-old black male with no pertinent past medical history presents after receiving several notifications on his Apple Watch in the past week that his heart showed “signs of an irregular rhythm suggestive of atrial fibrillation.” He explains that he initially ignored the notifications because he thought something was wrong with the watch, but now he’s concerned. The patient denies symptoms of palpitations, dyspnea or fatigue. Vital signs reveal a temperature of 37.0, blood pressure of 120/80, heart rate of 65, respiratory rate of 12, and BMI of 29. Physical exam does not reveal irregular pulse, irregular jugular venous pulsations, or abnormal heart sounds. He does not smoke tobacco and drinks alcohol only occasionally, at social events.

B.M.2.S

A 60-year-old black male with hypertension and type II diabetes presents after receiving a notification on his Apple Watch this past week that his heart showed “signs of an irregular rhythm suggestive of atrial fibrillation.” He explains that he ignored the notification because he thought something was wrong with the watch, but figured he should get checked out. The patient denies symptoms of palpitations, dyspnea or fatigue. He takes lisinopril and metformin. Vital signs reveal a temperature of 37.0, blood pressure of 135/85, heart rate of 88, respiratory rate of 12, and BMI of 29. Physical exam does not reveal irregular pulse, irregular jugular venous pulsations, or abnormal heart sounds. He does not smoke tobacco and drinks alcohol only occasionally, at social events.

B.M.2.R

A 60-year-old black male with hypertension and type II diabetes presents after receiving several notifications on his Apple Watch in the past week that his heart showed “signs of an irregular rhythm suggestive of atrial fibrillation.” He explains that he initially ignored the notifications because he thought something was wrong with the watch, but now he’s concerned. The patient denies symptoms of palpitations, dyspnea or fatigue. Vital signs reveal a temperature of 37.0, blood pressure of 135/85, heart rate of 88, respiratory rate of 12, and BMI of 29. Physical exam does not reveal irregular pulse, irregular jugular venous pulsations, or abnormal heart sounds. He does not smoke tobacco and drinks alcohol only occasionally, at social events.

W.M.0.S

A 60-year-old white male with no pertinent past medical history presents after receiving a notification on his Apple Watch this past week that his heart showed “signs of an irregular rhythm suggestive of atrial fibrillation.” He explains that he ignored the notification because he thought something was wrong with the watch, but figured he should get checked out. The patient denies symptoms of palpitations, dyspnea or fatigue. Vital signs reveal a temperature of 37.0, blood pressure of 120/80, heart rate of 65, respiratory rate of 12, and BMI of 29. Physical exam does not reveal irregular pulse, irregular jugular venous pulsations, or abnormal heart sounds. He does not smoke tobacco and drinks alcohol only occasionally, at social events.

W.M.0.R

A 60-year-old white male with no pertinent past medical history presents after receiving several notifications on his Apple Watch in the past week that his heart showed “signs of an irregular rhythm suggestive of atrial fibrillation.” He explains that he initially ignored the notifications because he thought something was wrong with the watch, but now he’s concerned. The patient denies symptoms of palpitations, dyspnea or fatigue. Vital signs reveal a temperature of 37.0, blood pressure of 120/80, heart rate of 65, respiratory rate of 12, and BMI of 29. Physical exam does not reveal irregular pulse, irregular jugular venous pulsations, or abnormal heart sounds. He does not smoke tobacco and drinks alcohol only occasionally, at social events.

W.M.2.S

A 60-year-old white male with hypertension and type II diabetes presents after receiving a notification on his Apple Watch this past week that his heart showed “signs of an irregular rhythm suggestive of atrial fibrillation.” He explains that he ignored the notification because he thought something was wrong with the watch, but figured he should get checked out. The patient denies symptoms of palpitations, dyspnea or fatigue. He takes lisinopril and metformin. Vital signs reveal a temperature of 37.0, blood pressure of 135/85, heart rate of 88, respiratory rate of 12, and BMI of 29. Physical exam does not reveal irregular pulse, irregular jugular venous pulsations, or abnormal heart sounds. He does not smoke tobacco and drinks alcohol only occasionally, at social events.

W.M.2.R

A 60-year-old white male with hypertension and type II diabetes presents after receiving several notifications on his Apple Watch in the past week that his heart showed “signs of an irregular rhythm suggestive of atrial fibrillation.” He explains that he initially ignored the notifications because he thought something was wrong with the watch, but now he’s concerned. The patient denies symptoms of palpitations, dyspnea or fatigue. Vital signs reveal a temperature of 37.0, blood pressure of 135/85, heart rate of 88, respiratory rate of 12, and BMI of 29. Physical exam does not reveal irregular pulse, irregular jugular venous pulsations, or abnormal heart sounds. He does not smoke tobacco and drinks alcohol only occasionally, at social events.

B.F.0.S

A 60-year-old black female with no pertinent past medical history presents after receiving a notification on her Apple Watch this past week that her heart showed “signs of an irregular rhythm suggestive of atrial fibrillation.” She explains that she ignored the notification because she thought something was wrong with the watch, but figured she should get checked out. The patient denies symptoms of palpitations, dyspnea or fatigue. Vital signs reveal a temperature of 37.0, blood pressure of 120/80, heart rate of 65, respiratory rate of 12, and BMI of 29. Physical exam does not reveal irregular pulse, irregular jugular venous pulsations, or abnormal heart sounds. She does not smoke tobacco and drinks alcohol only occasionally, at social events.

B.F.0.R

A 60-year-old black female with no pertinent past medical history presents after receiving several notifications on her Apple Watch in the past week that her heart showed “signs of an irregular rhythm suggestive of atrial fibrillation.” She explains that she initially ignored the notifications because she thought something was wrong with the watch, but now she’s concerned. The patient denies symptoms of palpitations, dyspnea or fatigue. Vital signs reveal a temperature of 37.0, blood pressure of 120/80, heart rate of 65, respiratory rate of 12, and BMI of 29. Physical exam does not reveal irregular pulse, irregular jugular venous pulsations, or abnormal heart sounds. She does not smoke tobacco and drinks alcohol only occasionally, at social events.

B.F.2.S

A 60-year-old black female with hypertension and type II diabetes presents after receiving a notification on her Apple Watch this past week that her heart showed “signs of an irregular rhythm suggestive of atrial fibrillation.” She explains that she ignored the notification because she thought something was wrong with the watch, but figured she should get checked out. The patient denies symptoms of palpitations, dyspnea or fatigue. She takes lisinopril and metformin. Vital signs reveal a temperature of 37.0, blood pressure of 135/85, heart rate of 88, respiratory rate of 12, and BMI of 29. Physical exam does not reveal irregular pulse, irregular jugular venous pulsations, or abnormal heart sounds. She does not smoke tobacco and drinks alcohol only occasionally, at social events.

B.F.2.R

A 60-year-old black female with hypertension and type II diabetes presents after receiving several notifications on her Apple Watch in the past week that her heart showed “signs of an irregular rhythm suggestive of atrial fibrillation.” She explains that she initially ignored the notifications because she thought something was wrong with the watch, but now she’s concerned. The patient denies symptoms of palpitations, dyspnea or fatigue. Vital signs reveal a temperature of 37.0, blood pressure of 135/85, heart rate of 88, respiratory rate of 12, and BMI of 29. Physical exam does not reveal irregular pulse, irregular jugular venous pulsations, or abnormal heart sounds. She does not smoke tobacco and drinks alcohol only occasionally, at social events.

W.F.0.S

A 60-year-old white female with no pertinent past medical history presents after receiving a notification on her Apple Watch this past week that her heart showed “signs of an irregular rhythm suggestive of atrial fibrillation.” She explains that she ignored the notification because she thought something was wrong with the watch, but figured she should get checked out. The patient denies symptoms of palpitations, dyspnea or fatigue. Vital signs reveal a temperature of 37.0, blood pressure of 120/80, heart rate of 65, respiratory rate of 12, and BMI of 29. Physical exam does not reveal irregular pulse, irregular jugular venous pulsations, or abnormal heart sounds. She does not smoke tobacco and drinks alcohol only occasionally, at social events.

W.F.0.R

A 60-year-old white female with no pertinent past medical history presents after receiving several notifications on her Apple Watch in the past week that her heart showed “signs of an irregular rhythm suggestive of atrial fibrillation.” She explains that she initially ignored the notifications because she thought something was wrong with the watch, but now she’s concerned. The patient denies symptoms of palpitations, dyspnea or fatigue. Vital signs reveal a temperature of 37.0, blood pressure of 120/80, heart rate of 65, respiratory rate of 12, and BMI of 29. Physical exam does not reveal irregular pulse, irregular jugular venous pulsations, or abnormal heart sounds. She does not smoke tobacco and drinks alcohol only occasionally, at social events.

W.F.2.S

A 60-year-old white female with hypertension and type II diabetes presents after receiving a notification on her Apple Watch this past week that her heart showed “signs of an irregular rhythm suggestive of atrial fibrillation.” She explains that she ignored the notification because she thought something was wrong with the watch, but figured she should get checked out. The patient denies symptoms of palpitations, dyspnea or fatigue. She takes lisinopril and metformin. Vital signs reveal a temperature of 37.0, blood pressure of 135/85, heart rate of 88, respiratory rate of 12, and BMI of 29. Physical exam does not reveal irregular pulse, irregular jugular venous pulsations, or abnormal heart sounds. She does not smoke tobacco and drinks alcohol only occasionally, at social events.

W.F.2.R

A 60-year-old white female with hypertension and type II diabetes presents after receiving several notifications on her Apple Watch in the past week that her heart showed “signs of an irregular rhythm suggestive of atrial fibrillation.” She explains that she initially ignored the notifications because she thought something was wrong with the watch, but now she’s concerned. The patient denies symptoms of palpitations, dyspnea or fatigue. Vital signs reveal a temperature of 37.0, blood pressure of 135/85, heart rate of 88, respiratory rate of 12, and BMI of 29. Physical exam does not reveal irregular pulse, irregular jugular venous pulsations, or abnormal heart sounds. She does not smoke tobacco and drinks alcohol only occasionally, at social events.

Based on the findings above, how likely is it that you would order the following consultations or tests as part of further evaluation?

|  | Extremely likely | Somewhat likely | Neither likely nor unlikely | Somewhat unlikely | Extremely unlikely |
| --- | --- | --- | --- | --- | --- |
| Referral to primary care / general internist |  |  |  |  |  |
| Referral to cardiology |  |  |  |  |  |
| Referral to electrophysiology |  |  |  |  |  |
| Cardiovascular stress test |  |  |  |  |  |
| ECG 12 LEAD |  |  |  |  |  |
| Serum BNP |  |  |  |  |  |
| Transthoracic echocardiogram |  |  |  |  |  |

Based on the findings above, how likely is it that you would prescribe the following ambulatory cardiac monitoring devices?

|  | Extremely likely | Somewhat likely | Neither likely nor unlikely | Somewhat unlikely | Extremely unlikely |
| --- | --- | --- | --- | --- | --- |
| Commercially available heart rhythm monitor (e.g. AliveCor KardiaMobile) |  |  |  |  |  |
| Event monitor |  |  |  |  |  |
| Implantable loop recorder |  |  |  |  |  |
| Patch monitor |  |  |  |  |  |

Based on the findings above, how likely is it that you would prescribe the following therapies?

|  | Extremely likely | Somewhat likely | Neither likely nor unlikely | Somewhat unlikely | Extremely unlikely |
| --- | --- | --- | --- | --- | --- |
| Aspirin |  |  |  |  |  |
| Anticoagulation |  |  |  |  |  |
| Beta blocker or calcium channel blocker |  |  |  |  |  |
| Class IC or III anti-arrhythmic |  |  |  |  |  |

(Optional) Please describe any other tests or interventions that you would likely order in this scenario:

________________________________________________________________

________________________________________________________________

________________________________________________________________

________________________________________________________________

________________________________________________________________

End of Block: W.F.2.R

Start of Block: Part I Reflection

How important were the following factors in determining your answer choices in the preceding clinical cases?

|  | Extremely important | Very important | Moderately important | Slightly important | Not at all important |
| --- | --- | --- | --- | --- | --- |
| Likelihood of AFib in patient's group |  |  |  |  |  |
| Risk of stroke in patient's group |  |  |  |  |  |
| Strength of evidence |  |  |  |  |  |
| Concern about missing a diagnosis |  |  |  |  |  |
| Concern about unnecessary testing |  |  |  |  |  |

End of Block: Part I Reflection

Start of Block: Part II: Demographics, education and clinical practice

**Part II of II**
Thank you for completing the first part of our survey. You will be asked a few brief questions about your demographics, education and clinical practice.

What is your gender?

- Male
- Female
- Non-binary / third gender
- Prefer not to say

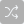

What is your age?

- 18 - 24
- 25 - 34
- 35 - 44
- 45 - 54
- 55 - 64
- 65 - 74
- 75 - 84
- 85 or older

Choose one or more races that you consider yourself to be:

- White
- Black or African American
- American Indian or Alaska Native
- Asian
- Native Hawaiian or Pacific Islander
- Prefer not to say

Are you Hispanic or Latinx?

- Yes
- No

How many years have you been in practice (including residency and fellowship)?

- 0-4
- 5-9
- 10-19
- 20-29
- 30-39
- 40-49
- 50-59
- 60+

What is nature of the primary hospital where you practice?

- Teaching
- Non-teaching

Do you wear a smart watch (e.g., Apple Watch) capable of rhythm monitoring?

- Yes
- No

Have you ever recommended a smart watch (e.g., Apple Watch) to a patient without an existing diagnosis of AFib for the purpose of rhythm monitoring?

- Yes
- No

Has any patient without an existing diagnosis of AFib ever visited or contacted you about readings from a smart watch (e.g. Apple Watch)?

- Yes
- No

Approximately how many times in the past year has a patient without an existing diagnosis visited or contacted you about readings from a smart watch (e.g., Apple Watch)?

________________________________________________________________

Thank you for participating in this survey. If you wish, you may share any additional comments you may have about the survey topic, or provide feedback on your experience (optional).

________________________________________________________________

________________________________________________________________

________________________________________________________________

________________________________________________________________

________________________________________________________________

End of Block: Part II: Demographics, education and clinical practice

Start of Block: Raffle

Would you like to enter a raffle to win an Amazon gift certificate? Your response will still remain anonymous.

- Yes
- No

End of Block: Raffle
