## Supplement 3 for "Physician Responses to Apple Watch-Detected Irregular Rhythm Alerts: A Case-Based Survey"

**Supplementary Table 1: Factors influencing respondents’ answers to cases.** Respondents were asked to rate the importance of the following factors in determining their answers to the cases. Data were missing for 6 respondents (6.3% overall).

|  | **Overall (N=95)** |
| --- | --- |
| **Likelihood of AFib in patient's group** |  |
| Extremely important | 23 (24.2%) |
| Very important | 25 (26.3%) |
| Moderately important | 26 (27.4%) |
| Slightly important | 10 (10.5%) |
| Not at all important | 5 (5.3%) |
| **Risk of stroke in patient's group** |  |
| Extremely important | 17 (17.9%) |
| Very important | 23 (24.2%) |
| Moderately important | 28 (29.5%) |
| Slightly important | 15 (15.8%) |
| Not at all important | 6 (6.3%) |
| **Strength of evidence** |  |
| Extremely important | 20 (21.1%) |
| Very important | 34 (35.8%) |
| Moderately important | 24 (25.3%) |
| Slightly important | 6 (6.3%) |
| Not at all important | 5 (5.3%) |
| **Concern about missing a diagnosis** |  |
| Extremely important | 11 (11.6%) |
| Very important | 30 (31.6%) |
| Moderately important | 26 (27.4%) |
| Slightly important | 16 (16.8%) |
| Not at all important | 6 (6.3%) |
| **Concern about unnecessary testing** |  |
| Extremely important | 14 (14.7%) |
| Very important | 32 (33.7%) |
| Moderately important | 29 (30.5%) |
| Slightly important | 10 (10.5%) |
| Not at all important | 4 (4.2%) |

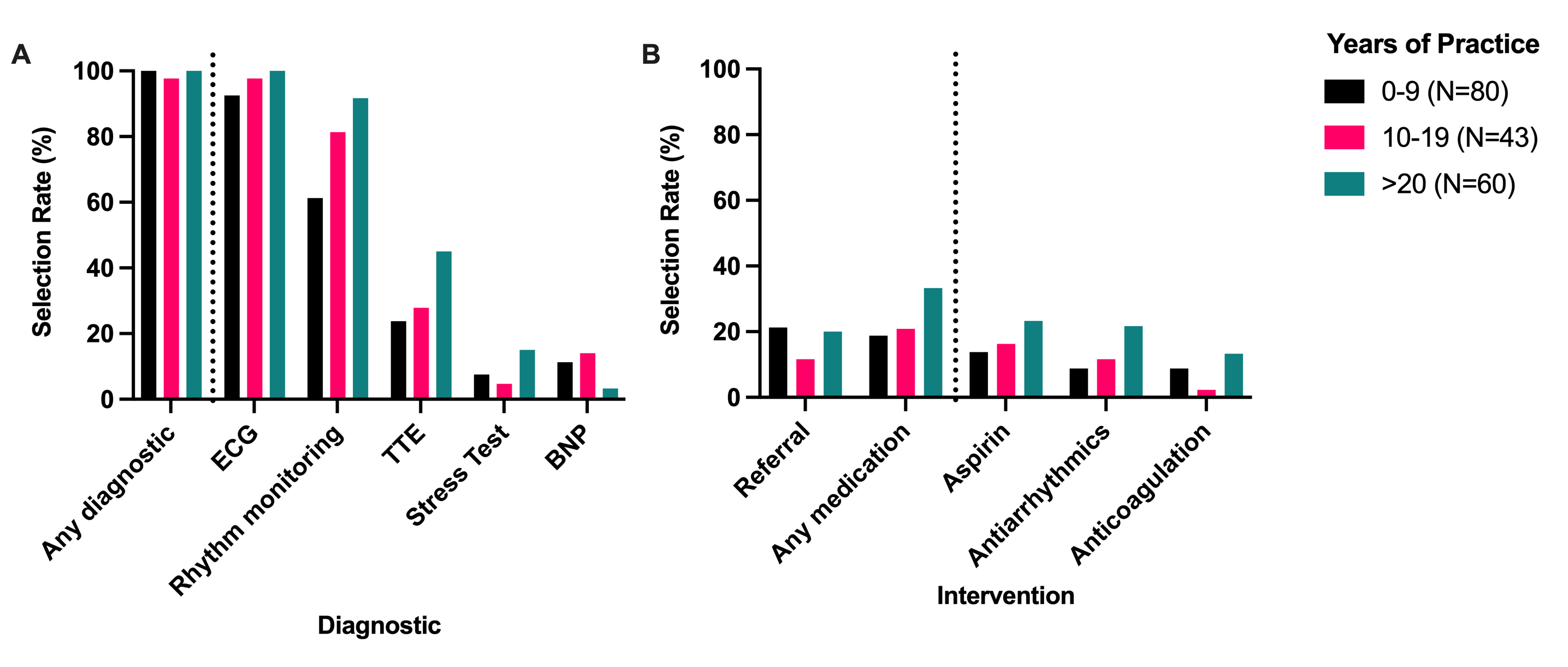

**Supplementary Figure 1: Rates of testing and intervention by respondent years in practice.**

**Supplementary Table 2: Rates of testing and intervention by respondent experience with smartwatches.** * 1 subject had missing data.

|  | **Wears a Smartwatch** | | | | | **Has Treated a Patient Who Reported a Wearable Alert** | | | | | | | | | | **Has Recommended Wearables to Patients** | | | | | | | |
| --- | --- | --- | --- | --- | --- | --- | --- | --- | --- | --- | --- | --- | --- | --- | --- | --- | --- | --- | --- | --- | --- | --- | --- |
|  | **Yes (N=56)** | **No (N=127)** | **P-value** | | | | **Yes (N=117)** | | | **No (N=66)** | | | **P-value** | | | | **Yes (N=42)** | | | **No (N=141)** | | | **P-value** |
| **Referral (PCP <> Cardiology/EP)** |  |  |  | | | |  | | |  | | |  | | | |  | | |  | | |  |
| Yes | 13 (23.2%) | 21 (16.5%) | 0.387 | | | | 22 (18.8%) | | | 12 (18.2%) | | | 1 | | | | 4 (9.5%) | | | 30 (21.3%) | | | 0.135 |
| No | 43 (76.8%) | 106 (83.5%) |  | | | | 95 (81.2%) | | | 54 (81.8%) | | |  | | | | 38 (90.5%) | | | 111 (78.7%) | | |  |
| **Any diagnostic test** |  |  |  | | | |  | | |  | | |  | | | |  | | |  | | |  |
| Yes | 56 (100%) | 126 (99.2%) | 1 | | | | 116 (99.1%) | | | 66 (100%) | | | 1 | | | | 42 (100%) | | | 140 (99.3%) | | | 1 |
| No | 0 (0%) | 1 (0.8%) |  | | | | 1 (0.9%) | | | 0 (0%) | | |  | | | | 0 (0%) | | | 1 (0.7%) | | |  |
| **Stress Test** |  |  |  | | | |  | | |  | | |  | | | |  | | |  | | |  |
| Yes | 7 (12.5%) | 10 (7.9%) | 0.484 | | | | 8 (6.8%) | | | 9 (13.6%) | | | 0.216 | | | | 5 (11.9%) | | | 12 (8.5%) | | | 0.727 |
| No* | 49 (87.5%) | 117 (92.1%) |  | | | | 109 (93.2%) | | | 57 (86.4%) | | |  | | | | 37 (88.1%) | | | 129 (91.5%) | | |  |
| **Electrocardiogram** |  |  |  | | | |  | | |  | | |  | | | |  | | |  | | |  |
| Yes | 54 (96.4%) | 122 (96.1%) | 1 | | | | 112 (95.7%) | | | 64 (97.0%) | | | 0.984 | | | | 40 (95.2%) | | | 136 (96.5%) | | | 1 |
| No | 2 (3.6%) | 5 (3.9%) |  | | | | 5 (4.3%) | | | 2 (3.0%) | | |  | | | | 2 (4.8%) | | | 5 (3.5%) | | |  |
| **Brain Natriuretic Peptide** |  |  |  | | | |  | | |  | | |  | | | |  | | |  | | |  |
| Yes | 5 (8.9%) | 12 (9.4%) | 1 | | | | 13 (11.1%) | | | 4 (6.1%) | | | 0.387 | | | | 2 (4.8%) | | | 15 (10.6%) | | | 0.396 |
| No | 51 (91.1%) | 115 (90.6%) |  | | | | 104 (88.9%) | | | 62 (93.9%) | | |  | | | | 40 (95.2%) | | | 126 (89.4%) | | |  |
| **Transthoracic echocardiogram** |  |  |  | | | |  | | |  | | |  | | | |  | | |  | | |  |
| Yes | 21 (37.5%) | 37 (29.1%) | 0.343 | | | | 41 (35.0%) | | | 17 (25.8%) | | | 0.258 | | | | 23 (54.8%) | | | 35 (24.8%) | | | <0.001 |
| No | 35 (62.5%) | 90 (70.9%) |  | | | | 76 (65.0%) | | | 49 (74.2%) | | |  | | | | 19 (45.2%) | | | 106 (75.2%) | | |  |
| **Ambulatory rhythm monitoring** |  |  |  | | | |  | | |  | | |  | | | |  | | |  | | |  |
| Yes | 43 (76.8%) | 96 (75.6%) | 1 | | | | 89 (76.1%) | | | 50 (75.8%) | | | 1 | | | | 40 (95.2%) | | | 99 (70.2%) | | | 0.002 |
| No | 13 (23.2%) | 31 (24.4%) |  | | | | 28 (23.9%) | | | 16 (24.2%) | | |  | | | | 2 (4.8%) | | | 42 (29.8%) | | |  |
| **Any medication** |  |  |  | | | |  | | |  | | |  | | | |  | | |  | | |  |
| Yes | 15 (26.8%) | 29 (22.8%) | 0.698 | | | | 33 (28.2%) | | | 11 (16.7%) | | | 0.116 | | | | 12 (28.6%) | | | 32 (22.7%) | | | 0.564 |
| No | 41 (73.2%) | 98 (77.2%) |  | | | | 84 (71.8%) | | | 55 (83.3%) | | |  | | | | 30 (71.4%) | | | 109 (77.3%) | | |  |
| **Aspirin** |  |  |  | | | |  | | |  | | |  | | | |  | | |  | | |  |
| Yes | 12 (21.4%) | 20 (15.7%) | 0.485 | | | | 23 (19.7%) | | | 9 (13.6%) | | | 0.394 | | | | 7 (16.7%) | | | 25 (17.7%) | | | 1 |
| No* | 44 (78.6%) | 107 (84.3%) |  | | | | 94 (80.3%) | | | 57 (86.4%) | | |  | | | | 35 (83.4%) | | | 116 (82.3%) | | |  |
| **Anticoagulation** |  |  |  | | | |  | | |  | | |  | | | |  | | |  | | |  |
| Yes | 6 (10.7%) | 10 (7.9%) | 0.732 | | | | 13 (11.1%) | | | 3 (4.5%) | | | 0.216 | | | | 4 (9.5%) | | | 12 (8.5%) | | | 1 |
| No | 50 (89.3%) | 117 (92.1%) |  | | | | 104 (88.9%) | | | 63 (95.5%) | | |  | | | | 38 (90.5%) | | | 129 (91.5%) | | |  |
| **Antiarrhythmics** |  |  |  | | | |  | | |  | | |  | | | |  | | |  | | |  |
| Yes | 8 (14.3%) | 17 (13.4%) | 1 | | | | 18 (15.4%) | | | 7 (10.6%) | | | 0.497 | | | | 6 (14.3%) | | | 19 (13.5%) | | | 1 |
| No | 48 (85.7%) | 110 (86.6%) |  | | | | 99 (84.6%) | | | 59 (89.4%) | | |  | | | | 36 (85.7%) | | | 122 (86.5%) | | |  |

**Supplementary Table 3. Distribution of case characteristics.**

|  | **Overall (N=192)** |
| --- | --- |
| **Race** |  |
| Black | 94 (49.0%) |
| White | 98 (51.0%) |
| **Sex** |  |
| Female | 100 (52.1%) |
| Male | 92 (47.9%) |
| **Stroke Risk Factors (Hypertension & Type 2 Diabetes)** |  |
| No | 100 (52.1%) |
| Yes | 92 (47.9%) |
| **Number of Notifications** |  |
| Multiple | 100 (52.1%) |
| One | 92 (47.9%) |

**Supplementary Table 4: Rates of testing and intervention by hypothetical patient characteristics.** * 1 subject had missing data.

|  | **Sex** | | | | | **Stroke Risk Factors**  **(Hypertension & Type 2 Diabetes)** | | | | | **Number of Notifications** | | | | | |
| --- | --- | --- | --- | --- | --- | --- | --- | --- | --- | --- | --- | --- | --- | --- | --- | --- |
|  | **Male (N=92)** | **Female (N=100)** | **P-value** | | **No (N=100)** | | | **Yes (N=92)** | **P-value** | | | **One (N=92)** | | **Multiple (N=100)** | | **P-value** |
| **Referral (PCP <> Cardiology/EP)** |  |  |  | |  | | |  |  | | |  | |  | |  |
| Yes | 15 (16.3%) | 21 (21.0%) | 0.496 | | 18 (18.0%) | | | 18 (19.6%) | 0.897 | | | 14 (15.2%) | | 22 (22.0%) | | 0.293 |
| No | 77 (83.7%) | 78 (78.0%) |  | | 82 (82.0%) | | | 73 (79.3%) |  | | | 78 (84.8%) | | 77 (77.0%) | |  |
| Missing | 0 (0%) | 1 (1.0%) |  | | 0 (0%) | | | 1 (1.1%) |  | | | 0 (0%) | | 1 (1.0%) | |  |
| **Any diagnostic test** |  |  |  | |  | | |  |  | | |  | |  | |  |
| Yes | 92 (100%) | 99 (99.0%) |  | | 99 (99.0%) | | | 92 (100%) | 1 | | | 91 (98.9%) | | 100 (100%) | | 0.967 |
| No | 0 (0%) | 1 (1.0%) | 1 | | 1 (1.0%) | | | 0 (0%) |  | | | 1 (1.1%) | | 0 (0%) | |  |
| **Stress Test** |  |  |  | |  | | |  |  | | |  | |  | |  |
| Yes | 10 (10.9%) | 9 (9.0%) |  | | 7 (7.0%) | | | 12 (13.0%) | 0.256 | | | 9 (9.8%) | | 10 (10.0%) | | 1 |
| No* | 82 (89.1%) | 91 (91.0%) | 0.866 | | 93 (93.0%) | | | 80 (87.0%) |  | | | 83 (90.2%) | | 90 (90.0%) | |  |
| **Electrocardiogram** |  |  |  | |  | | |  |  | | |  | |  | |  |
| Yes | 90 (97.8%) | 95 (95.0%) |  | | 96 (96.0%) | | | 89 (96.7%) | 1 | | | 90 (97.8%) | | 95 (95.0%) | | 0.51 |
| No | 2 (2.2%) | 5 (5.0%) | 0.51 | | 4 (4.0%) | | | 3 (3.3%) |  | | | 2 (2.2%) | | 5 (5.0%) | |  |
| **Brain Natriuretic Peptide** |  |  |  | |  | | |  |  | | |  | |  | |  |
| Yes | 13 (14.1%) | 4 (4.0%) |  | | 8 (8.0%) | | | 9 (9.8%) | 0.857 | | | 10 (10.9%) | | 7 (7.0%) | | 0.491 |
| No | 79 (85.9%) | 96 (96.0%) | 0.0268 | | 92 (92.0%) | | | 83 (90.2%) |  | | | 82 (89.1%) | | 93 (93.0%) | |  |
| **Transthoracic echocardiogram** |  |  |  | |  | | |  |  | | |  | |  | |  |
| Yes | 33 (35.9%) | 30 (30.0%) |  | | 28 (28.0%) | | | 35 (38.0%) | 0.185 | | | 32 (34.8%) | | 31 (31.0%) | | 0.686 |
| No | 59 (64.1%) | 70 (70.0%) | 0.477 | | 72 (72.0%) | | | 57 (62.0%) |  | | | 60 (65.2%) | | 69 (69.0%) | |  |
| **Ambulatory rhythm monitoring** |  |  |  | |  | | |  |  | | |  | |  | |  |
| Yes | 70 (76.1%) | 78 (78.0%) |  | | 77 (77.0%) | | | 71 (77.2%) | 1 | | | 70 (76.1%) | | 78 (78.0%) | | 0.886 |
| No | 22 (23.9%) | 22 (22.0%) | 0.886 | | 23 (23.0%) | | | 21 (22.8%) |  | | | 22 (23.9%) | | 22 (22.0%) | |  |
| **Any medication** |  |  |  | |  | | |  |  | | |  | |  | |  |
| Yes | 22 (23.9%) | 26 (26.0%) |  | | 21 (21.0%) | | | 27 (29.3%) | 0.243 | | | 24 (26.1%) | | 24 (24.0%) | | 0.868 |
| No | 70 (76.1%) | 74 (74.0%) | 0.868 | | 79 (79.0%) | | | 65 (70.7%) |  | | | 68 (73.9%) | | 76 (76.0%) | |  |
| **Aspirin** |  |  |  | |  | | |  |  | | |  | |  | |  |
| Yes | 16 (17.4%) | 19 (19.0%) |  | | 16 (16.0%) | | | 19 (20.7%) | 0.539 | | | 16 (17.4%) | | 19 (19.0%) | | 0.948 |
| No* | 76 (82.6%) | 81 (81.0%) | 0.893 | | 84 (84.0%) | | | 73 (79.3%) |  | | | 76 (82.6%) | | 81 (81.0%) | |  |
| **Anticoagulation** |  |  |  | |  | | |  |  | | |  | |  | |  |
| Yes | 9 (9.8%) | 9 (9.0%) |  | | 7 (7.0%) | | | 11 (12.0%) | 0.353 | | | 9 (9.8%) | | 9 (9.0%) | | 1 |
| No | 83 (90.2%) | 91 (91.0%) | 1 | | 93 (93.0%) | | | 81 (88.0%) |  | | | 83 (90.2%) | | 91 (91.0%) | |  |
| **Antiarrhythmics** |  |  |  | |  | | |  |  | | |  | |  | |  |
| Yes | 14 (15.2%) | 15 (15.0%) |  | | 13 (13.0%) | | | 16 (17.4%) | 0.518 | | | 16 (17.4%) | | 13 (13.0%) | | 0.518 |
| No | 78 (84.8%) | 85 (85.0%) | 1 | | 87 (87.0%) | | | 76 (82.6%) |  | | | 76 (82.6%) | | 87 (87.0%) | |  |
